# Ultrarare Variants in Genes Involved in Intestinal Microbiota and Permeability Homeostasis in Youth with Developmental and Neuropsychiatric Deteriorations

**DOI:** 10.64898/2026.05.29.26353976

**Authors:** Jennifer Frankovich, Robert A. Dubin, Chetna Natarajan, Noelle Schlenk, Erika Pedrosa, Erin Stolte, Nicole Rice, Anjana Soorajkumar, Dhanya Vettiatil, Peter J. van der Spek, Janet L. Cunningham, Herbert M. Lachman

## Abstract

Abnormalities in the gut microbiome, intestinal permeability, and the gut-immune-brain axis are increasingly linked to neuropsychiatric disorders, neurodegenerative disorders, inflammatory bowel disease (IBD), and other immunologic/autoimmune conditions. We investigated these phenomena in 128 youth with Pediatric Acute-Onset Neuropsychiatric Syndrome (PANS) and individuals with autism spectrum disorder (ASD) and other neurodevelopmental disorders (NDD) characterized by profound, unexplained deteriorations/regressions in developmental, neuropsychiatric, and behavioral functioning. Previous studies we have carried out showed that immune dysregulation and DNA damage response (DDR) gene mutations are implicated in a subset of these patients. The current study examines the role of genetic variants affecting intestinal homeostasis. We report a series of patients exhibiting both neuropsychiatric deterioration and gastrointestinal symptoms. Genetic analysis identified ultrarare (minor allele frequency < 0.001) pathogenic or likely pathogenic variants in eight genes primarily expressed in the intestines and associated with IBD, dysbiosis, or intestinal permeability. Across thirteen patients, mutations were identified in *DUOX2* (n=4), *SLC10A2* (n=2), *UNC45A*, *TTC7A*, *LGALS4*, *SI*, *CCR9*, *MEP1B,* and *BACH2*. While these findings suggest a potential role for genetic variants governing intestinal homeostasis in these cases of neuropsychiatric decline, their presence in only a small subgroup necessitates larger, prospective cohorts to determine whether these variants are statistically significant and play a definitive role in the pathogenesis of these disorders.

## Introduction

Pediatric Acute-Onset Neuropsychiatric Syndrome (PANS) is characterized by the core symptoms of abrupt onset obsessive-compulsive disorder (OCD) and/or restricted eating along with secondary symptoms: anxiety, emotional lability, irritability/rage, cognitive decline, sleep disturbance, sensory dysregulation, movement abnormalities, and urinary symptoms (new onset enuresis or urinary frequency) ^1–3^. While the diagnostic criteria require two secondary symptoms, most patients have five or more that start abruptly alongside OCD and/or restricted eating. The course is typically relapsing and remitting ^1, 4^ and is associated with high care-giver burden ^5^. PANS and other post-infectious/inflammation-related regressions can occur in children with premorbid ASD or other NDD ^6, 7^.

We previously identified pathogenic variants in genes that code for regulators of the DNA damage response (DDR) (e.g., *PPM1D, ATM, CHK2*, and several members of the Fanconi anemia complex) in patients with PANS and related deteriorations ^6, 8, 9^. We hypothesized that defects in DDR activate the innate immune system (e.g., cGAS-STING; AIM2 inflammasomes) contributing to neuroinflammation through activation of type I interferons and other innate immune pathways. Abnormalities in DDR can also disrupt gene expression and mitochondrial function, and cause senescence ^10–13^. De novo mutations in genes that affect interferon signaling have also been identified in Down Syndrome Regression Syndrome ^14^. In addition, pathogenic variants in chromatin-related genes, many of which have DNA repair functions, have been found in PANS and neuropsychiatric regression ^15^.

In the course of our screening, we also discovered a case with a pathogenic variant in the *DUOX2* gene. *DUOX2* is a compelling candidate gene as it encodes dual oxidase 2, an enzyme that plays a crucial role in producing hydrogen peroxide and thyroid hormone ^16, 17^. It is expressed in the apical membrane of gastrointestinal (GI) epithelia and contributes to IBD by disrupting microbiota-immune homeostasis ^18–21^. It also contributes to host defense and chronic inflammation, in part through interactions with IL-17 signaling, which is a regulator of gut barrier immunity ^22^. The identification of a second case with a likely pathogenic *DUOX2* mutation led to the purpose of this study where we reexamined the previously published cases, as well as an expanded cohort to identify pathogenic or likely pathogenic variants in genes expressed primarily in the gut that have been implicated in gut dysbiosis, intestinal permeability, and IBD. Clinical observations of intestinal symptoms in PANS and ASD suggest that gut dysbiosis, intestinal permeability, and/or gut-immune-brain axis signaling play a role in their pathogenesis, which warrants further investigation ^23–28^.

To the best of our knowledge, this exploratory study is the first to report specific genetic variants linking gut dysbiosis or intestinal permeability in ASD regression and PANS. Based on our findings, we hypothesize that increased susceptibility to acute neuropsychiatric decompensation following infections or non-infectious stressors may be driven by ultrarare genetic variants found in a small subset of PANS and ASD regression cases that contribute to a proinflammatory state by impacting the gut-immune-brain axis.

## Results

We analyzed WES data and reviewed genetic reports of 105 PANS and regression cases in ASD/NDD from Einstein cohort and i genetic reports in a second cohort of 23 patients with PANS from the Stanford Cohort. Thirteen cases (eight males and five females) were identified to have ultrarare variants in eight genes expressed exclusively or primarily in the intestines, which have also been implicated in IBD, intestinal dysbiosis, and/or intestinal permeability (*DUOX2, SLC10A2, TTC7A, LGALS4, SI, CCR9, MEP1B, UNC45A,* and *BACH2*.). An exception is *TTC7A*, which is expressed ubiquitously but shares a role in gut antimicrobial defense. 12 of 13 cases met the clinical criteria for PANS, of whom 2 had also had atypical neurodevelopment (ASD and developmental delay); one (case 13) was diagnosed with ASD and recurrent episodes of catatonia. Three cases (1, 8, 10) have mutations in DNA repair genes that were previously described ^6^. The additional mutations described in this study suggest a combination of genetic and molecular vulnerabilities. Two of the variants are de novo; five were transmitted by a parent, and in seven, transmission was not determined (**Table 1**). Six of the variants were classified as pathogenic or likely pathogenic by the Franklin database; three *DUOX2* variants (cases 2-4), *SLC10A2* (case 5)*, TTC7A* (case 7), and *SI* (case 9). Among the seven genes that were scored as variants of undetermined significance (VUS) by Franklin included a *DUOX2* variant with a predicted splice donor loss (case 1); a stop-gain variant in *LGALS4* with a high Combined Annotation Dependent Depletion (CADD) score of 39 (case 8; see methods); one of the *SLC10A* variants that was called pathogenic by AlphaMissense (AM) and had a high CADD score of 31 (case 6); a de novo frameshift variant in *CCR9* (case 10); a stop-gain variant in *MEP1B* with a CADD score of 36 (case 11); a nonsynonymous variant in *UNC45A* that was scored as pathogenic by AM that has a CADD score of 27 (case 12); and a novel de novo variant that causes a frameshift in *BACH2* (case 13). In addition to *DUOX2*, Case 4 also has an *UNC45A* variant that was scored as pathogenic by AM.

**Table 1:**
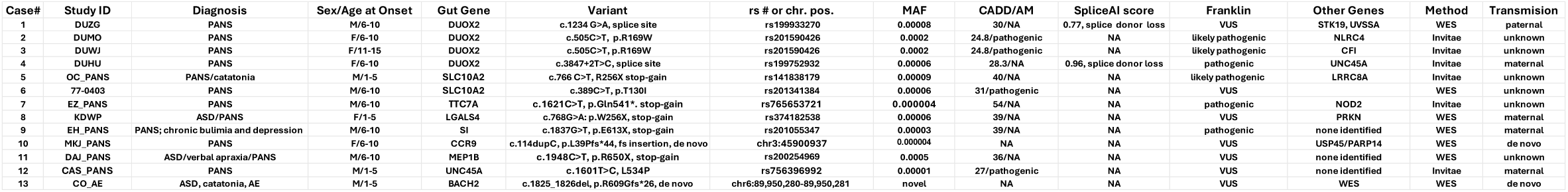
List of Variants: PANS (pediatric acute-onset neuropsychiatric syndrome) ASD (atism spectrum disorder); AE (autoimmune encephalopathy), AM (AlphaMissense); CADD (Combined Annotation Dependent Depletion) score; chr. pos. (chromosomal position) based on GRCh38/hg38 build; NA (not applicable); MAF (minor allele frequency)

### Cases 1-4: DUOX2

**Case 1 is** a typically developing child with recurrent infections, chronic abdominal pain, and diarrhea, and eventually met criteria for PANS who was found to have splice disrupting variants in two DNA repair genes *STK19* and *UVSSA* (see **Supplementary Table S1** for all secondary mutations) ^6^. He has also developed arthritis with inflammation affecting multiple vertebrae; arthritis (especially enthesitis) is found in nearly 30% of PANS patients ^29^. This patient also has a splice site mutation in *DUOX2,* which is predicted to disrupt a splice donor site (see **Table 1** for all primary gut gene variants). Both parents have a history of gluten intolerance, one of whom has been diagnosed with celiac disease. There are 1st and degree relatives with Hashimoto’s Thyroiditis and a sibling with PANS that is currently in remission (see **Table 2** for family history and summary of autoimmune disorders and gastrointestinal problems in cases). The sibling with PANS does not carry the *DUOX2*, *STK19* or *UVSSA* variants. Instead, the sibling has a common *NOD2* variant (MAF 0.02) found in case 7 that is associated with common variable immune deficiency (CVID), Blau syndrome and Yao syndrome in many studies ^30–34^.

**Table 2:**
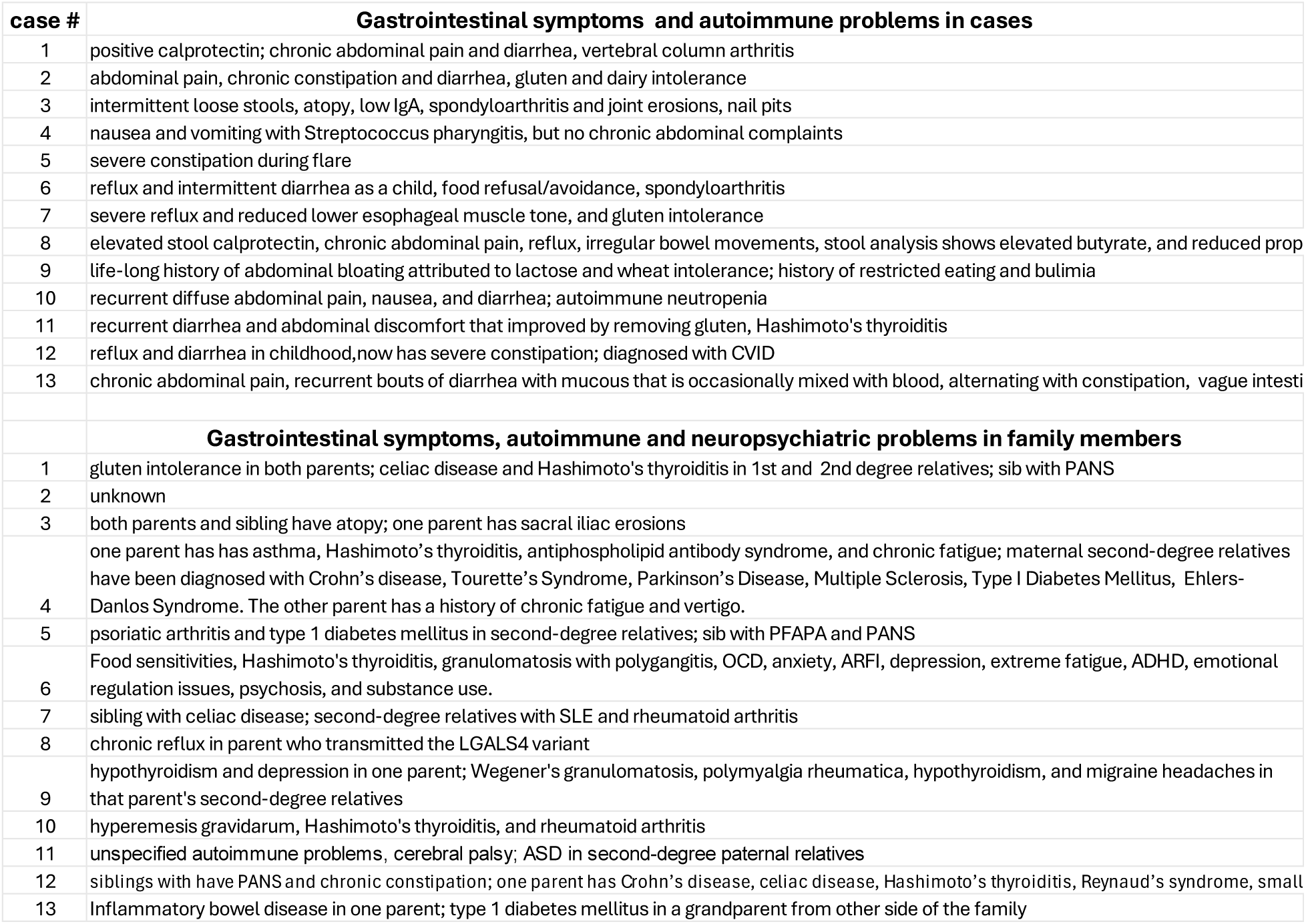
Gastrointestinal symptoms and autoimmune disorders in cases, and gastrointestinal symptoms, autoimmune disorders and neuropsychiatric disorders in family members.

| case # | Gastrointestinal symptoms and autoimmune problems in cases |
| --- | --- |
| 1 | positive calprotectin; chronic abdominal pain and diarrhea, vertebral column arthritis |
| 2 | abdominal pain, chronic constipation and diarrhea, gluten and dairy intolerance |
| 3 | intermittent loose stools, atopy, low IgA, spondyloarthritis and joint erosions, nail pits |
| 4 | nausea and vomiting with Streptococcus pharyngitis, but no chronic abdominal complaints |
| 5 | severe constipation during flare |
| 6 | reflux and intermittent diarrhea as a child, food refusal/avoidance, spondyloarthritis |
| 7 | severe reflux and reduced lower esophageal muscle tone, and gluten intolerance |
| 8 | elevated stool calprotectin, chronic abdominal pain, reflux, irregular bowel movements, stool analysis shows elevated butyrate, and reduced prop |
| 9 | life-long history of abdominal bloating attributed to lactose and wheat intolerance; history of restricted eating and bulimia |
| 10 | recurrent diffuse abdominal pain, nausea, and diarrhea; autoimmune neutropenia |
| 11 | recurrent diarrhea and abdominal discomfort that improved by removing gluten, Hashimoto's thyroiditis |
| 12 | reflux and diarrhea in childhood, now has severe constipation; diagnosed with CVID |
| 13 | chronic abdominal pain, recurrent bouts of diarrhea with mucous that is occasionally mixed with blood, alternating with constipation, vague intesti |
|  | **Gastrointestinal symptoms, autoimmune and neuropsychiatric problems in family members** |
| 1 | gluten intolerance in both parents; celiac disease and Hashimoto's thyroiditis in 1st and 2nd degree relatives; sib with PANS |
| 2 | unknown |
| 3 | both parents and sibling have atopy; one parent has sacral iliac erosions |
| 4 | one parent has has asthma, Hashimoto's thyroiditis, antiphospholipid antibody syndrome, and chronic fatigue; maternal second-degree relatives have been diagnosed with Crohn's disease, Tourette's Syndrome, Parkinson's Disease, Multiple Sclerosis, Type I Diabetes Mellitus, Ehlers-Danlos Syndrome. The other parent has a history of chronic fatigue and vertigo. |
| 5 | psoriatic arthritis and type 1 diabetes mellitus in second-degree relatives; sib with PFAPA and PANS |
| 6 | Food sensitivities, Hashimoto's thyroiditis, granulomatosis with polygangitis, OCD, anxiety, ARFI, depression, extreme fatigue, ADHD, emotional regulation issues, psychosis, and substance use. |
| 7 | sibling with celiac disease; second-degree relatives with SLE and rheumatoid arthritis |
| 8 | chronic reflux in parent who transmitted the LGALS4 variant |
| 9 | hypothyroidism and depression in one parent; Wegener's granulomatosis, polymyalgia rheumatica, hypothyroidism, and migraine headaches in that parent's second-degree relatives |
| 10 | hyperemesis gravidarum, Hashimoto's thyroiditis, and rheumatoid arthritis |
| 11 | unspecified autoimmune problems, cerebral palsy; ASD in second-degree paternal relatives |
| 12 | siblings with have PANS and chronic constipation; one parent has Crohn's disease, celiac disease, Hashimoto's thyroiditis, Reynaud's syndrome, small |
| 13 | Inflammatory bowel disease in one parent; type 1 diabetes mellitus in a grandparent from other side of the family |

**Case 2** is a typically developing child with chronic abdominal pain, constipation, diarrhea, dairy and gluten intolerance, diagnosed with PANS, who carries an ultrarare, likely pathogenic nonsynonymous mutation in *DUOX2*, which was confirmed by Sanger sequencing. There is also a history of recurrent constipation, diarrhea, and intolerance to dairy products. The child also has a novel variant in *NLRC4* that was scored as a VUS by Franklin but was ambiguous by AM. She was adopted and little is known about the history of the biological parents.

**Case 3** is a typically developing child with a history of intermittent loose stools, recurrent mouth sores, eczema, food allergies (tree nuts), allergic rhinitis, asthma, low immunoglobulins (IgG, IgA, IgM), extensive nail pits, suspected early spondyloarthritis vs psoriatic arthritis (erosions in temporomandibular joint and limited back flexion), and a history of relapsing/remitting PANS that is now persistent with severe OCD, suicidal thoughts, overwhelming anxiety, severe/persistent nightmares, and anhedonia. She presented in her fourth flare, which coincided with the rapid development of multiple dental cavities, complement cascade activation (low C3, high C3d, low C4), elevated ESR, high D-dimers, livedo reticularis, and leukopenia (prior to immunomodulation).

Both parents and a sibling have atopy, and one of the parents has erosions in joints including the sacroiliac joints and suspicion for early/mild spondylarthritis, and possible reactive arthritis vs gout. Transmission is not known but case 3 harbors the same *DUOX2* mutation found in case 2, despite a minor allele frequency of 0.0002; they are not related. She also has a likely pathogenic variant in CFI (complement factor I).

**Case 4** is a typically developing child diagnosed with relapsing/remitting post-Streptococcal PANS with flares spanning 10 years. Streptococcal pharyngitis is typically presented with GI symptoms. There is a strong history of autoimmune disorders in a parent and a history of Crohn’s disease in a second degree relative from the same parent. Case 4 has a splice-site mutation in *DUOX2* that is predicted to cause a splice donor loss. She also has an ultrarare variant in the *UNC45A* gene, which was scored as a VUS by Franklin, but is pathogenic according to AM. *UNC45A* codes for an HSP90 co-chaperone that regulates intestinal epithelial barrier integrity and repair ^35^. The parent with a history of Crohn’s disease transmitted the *DUOX2* and *UNC45A* variants. The *UNC45A* variant is different from the one found in case 12.

### Case 5: SLC10A2

Case 5 is a typically developing child who developed relapsing-remitting PANS and recurrent episodes of catatonia characterized by walking and eating refusal, poverty of speech, and obstipation. Flares were frequently associated with severe exacerbations of constipation. There is a history of psoriatic arthritis and type 1 diabetes mellitus in second-degree relatives. He also has a younger sibling with Periodic Fever, Aphthous Stomatitis, Pharyngitis, and Adenitis (PFAPA) who has had two PANS episodes, but no GI symptoms. This patient and his sibling were found to have a likely pathogenic stop-gain variant in the *SLC10A2* gene, which codes for the apical sodium-dependent bile acid transporter (ASBT). It is expressed almost exclusively in the small intestine and plays a key role in bile acid reabsorption (see discussion). The proband, but not his sibling, also carries an ultrarare *LRRC8A* variant that codes for a component of the volume-regulated anion channel (VRAC), which plays a role in amino acid transport, cell survival, and immune cell development ^36^. The *LRRC8A* variant was scored as a VUS by Franklin but is classified as pathogenic by AM.

### Case 6: SLC10A2

Case 6 is typically developing but had reflux and intermittent diarrhea, and longstanding separation anxiety, noise sensitivity, and sleep difficulties. Following a persistent low-grade fever, the patient experienced a subacute onset of intrusive thoughts, fear of vomiting, escalation of separation anxiety, decision-making difficulties, visual hypersensitivity, strong irritability and aggressive outburst. He had a subsequent relapse four months later (while under the care of J.F.) that was hyper-acute-onset and thus he met PANS criteria. He also had insidious onset of back pain which became severe/debilitating with eventual diagnosis of spondyloarthritis diagnosed at two academic centers. Back pain and neuropsychiatric symptoms eventually resolved while on sulfasalazine, low-dose methotrexate, and a TNF inhibitor alongside standard of care for his psychiatric symptoms. The transmission is not known. There is notable family history of thyroiditis, Avoidant/Restrictive Food Intake Disorder (ARFID), and depression one parent, including second-degree relatives from the same parent who have been diagnosed with depression, food sensitivity, recurrent strep, transient synovitis, granulomatosis with polyangiitis, and psychotic episodes. Second-degree relatives from the other parent have a notable history of depression, ADD, OCD, and substance use. The patient is heterozygous for a VUS in *SLC10A2* that has a CADD score of 31 and was scored as pathogenic by AM.

### Case 7: TTC7A

**Case 7** is typically developing but acquired relapsing-remitting PANS (with flares spanning a decade) followed by the diagnoses of POTS and hypermobility spectrum disorder (HSD). There is a history of severe reflux and reduced lower esophageal muscle tone and gluten intolerance (but not celiac disease). Family history is significant for a sibling who does not have PANS but carries the diagnosis of celiac disease, dysautonomia, HSD, and abdominal vascular compression syndrome. Second-degree relatives from one parent have been diagnosed with SLE and rheumatoid arthritis. Both case 7 and his sibling have a pathogenic nonsense mutation in *TTC7A*, which codes for, a scaffolding protein that maintains intestinal epithelial integrity and immune system homeostasis. Homozygosity or compound heterozygosity for loss-of-function mutations cause profound disruption of the epithelial barrier along the entire gastrointestinal tract, resulting in combined immunodeficiency with multiple intestinal atresia (CID-MIA), tricho-hepato-enteric syndrome (SD/THE), and IBD ^37–39^. Both siblings also carry the same *NOD2* common variant found in the sibling of case 1.

### Case 8: LGALS4

Case 8 has developmental delay along with non-specific intestinal symptoms, elevated stool calprotectin, and behavioral/psychiatric deterioration that met PANS criteria. She has a de novo *PRKN* exon 2/3 deletion that was described in our recent paper ^6^. The child also suffered from severe gastrointestinal reflux in the first year of life, resulting in poor weight gain and need for ongoing chronic acid suppression. She also has stop-gain mutation in *LGALS4* (Galectin-4). Although this variant was classified as a VUS by Franklin, the CADD score is very high; 39. This gene is expressed almost exclusively in epithelial cells in the transverse colon and terminal ileum. LGALS4 and other galectins are involved in the development of IBD by regulating T-cell function in the gut and affecting host-gut microbe interactions ^40, 41^. The parent who transmitted the *LGALS4* variant also has chronic reflux.

### Case 9: SI

Case 9 was typically developing and was diagnosed with PANS based on the abrupt onset of OCD, anxiety, mood disturbance, oppositional behavior, night terrors, and rage following an emotionally stressful situation at school and a Coxsackievirus infection. He had a persistent course punctuated by flares following infections. Although there was some improvement with selective serotonin reuptake inhibitors (SSRIs) and lithium, he never returned to baseline. Severe anxiety and depression with suicidal thoughts persisted into young adulthood, along with dysmorphia, restricted eating, and bulimia. There is also a history of gluten and dairy intolerance with recurrent bloating. The family history is positive for hypothyroidism and depression in first-degree relatives, and Wegener’s granulomatosis, polymyalgia rheumatica, hypothyroidism, and migraine headaches in second-degree relatives. WES identified a stop-gain mutation in the SI gene, which codes for the enzyme sucrose-isomaltose, that is expressed in the small intestine and is needed for the digestion of sucrose and maltose. SI deficiency is a cause of diarrhea-predominant irritable bowel syndrome ^42, 43^.

### Case 10: CCR9

Case 10 is a typically developing child with recurrent diffuse abdominal discomfort, diarrhea, and nausea (negative gastroenterology evaluation), autoimmune neutropenia, relapsing-remitting PANS and persistent sleep dysfunction, emotional outbursts, and oppositional behavior. There is a history of hyperemesis gravidarum, Hashimoto’s thyroiditis, and rheumatoid arthritis. A novel, de novo frameshift mutation was identified in *CCR9* (confirmed by Sanger sequencing), which codes for the CCL25 chemokine receptor. CCL25 regulates gut homeostasis and permeability ^44–46^. Pathogenic mutations were also identified in genes involved in DNA repair, including *USP45* and *PARP14,* which were described in our recent paper ^6^. PARP14 regulates innate immune function ^47^, and altered expression has been found in five different gene expressions datasets in Systemic Lupus Erythematosus (SLE) ^48^. Interestingly, Parp14 deficiency causes increased rectal bleeding in a mouse IBD model by impairing colon epithelial barrier integrity ^49^.

### Case 11: MEP1B

Case 11 was diagnosed with ASD, verbal apraxia, attention deficit hyperactivity disorder (ADHD), and relapsing-remitting PANS since early childhood. He also has a history of recurrent abdominal discomfort and diarrhea (improved by removing dietary gluten) and Hashimoto’s thyroiditis. There is a history of unspecified autoimmune problems, cerebral palsy, and ASD in second-degree paternal relatives. WES identified an ultrarare stop-gain mutation in *MEP1B* that is predicted to disrupt the membrane-binding domain (amino acids 653-673). The mutation was classified as a VUS by Franklin, but it has a high CADD score of 36. MEP1B (meprin A subunit beta) is a metalloprotease that plays a role in the hydrolysis of a variety of proteins that have been implicated in cancer, intestinal inflammation, and extracellular matrix (ECM) remodeling ^50^.

### Case 12: UNC45A

This is a boy who has had infection-induced neuropsychiatric symptoms since childhood that meet PANS criteria, which have persisted for more than 10 years. He was typically developing but was referred to a gastroenterologist as a child for reflux and diarrhea, which resolved by avoiding milk. He also had repeated bouts of otitis media, which necessitated the insertion of tympanostomy tubes, and was diagnosed with CVID. As a teenager, he has suffered from severe constipation in addition to neuropsychiatric symptoms, the latter of which have improved with IVIg. He has an ultrarare mutation in the *UNC45A* gene that was scored as a VUS by Franklin, but is pathogenic according to AM. In addition, the CADD score is 27. The variant is different from the one described in case 4. UNC45A codes for a protein that regulates intestinal epithelial barrier integrity and repair, as mentioned in case 4 ^35^. He has two younger siblings who both have chronic constipation and neuropsychiatric symptoms consistent with PANS. However, they have not been formally diagnosed. They also inherited the *UNC45A* variant, which was transmitted from a parent who has been diagnosed with Crohn’s disease, celiac disease, Hashimoto’s thyroiditis, Reynaud’s syndrome, small intestine bacterial overgrowth (SIBO), OCD, anxiety, and depression.

### Case 13: BACH2

Case 13 was diagnosed with non-verbal ASD and has had recurrent episodes of catatonia for three years that require ECT therapy several times a week. Development was typical until age 2.5 when there was an abrupt loss of language skills that progressed to catatonia. He has had many episodes of abrupt behavioral regressions, often preceded by infections, that would commonly progress to stuporous or excited catatonia.

The child also has severe recurrent bouts of diarrhea with mucus, occasionally mixed with blood, alternating with obstipation. Fecal calprotectin was elevated, but colonoscopy was negative. Severe gut symptoms often precede the decline to catatonia. Bowel symptoms and catatonia have responded well to sirolimus. A PET scan obtained during a regression episode showed some hypermetabolism of the left frontal and posterior parietal lobes compared with the right as well as hypermetabolism in the superior temporal gyrus.

The child has an immune deficiency characterized by recurrent infections, low IgG/IgM levels, near-absent B cells, and a poor response to childhood vaccines. One parent has been diagnosed with both UC and Crohn’s disease, and there is second degree relative from the other parent has type 1 diabetes mellitus. Family trio exome sequencing revealed a novel de novo frameshift mutation in *BACH2*, which codes for a transcription factor that plays a crucial role in regulating T cells and B cells ^51^. Heterozygous mutations in the *BACH2* gene can disrupt immune regulation and increase susceptibility to autoimmune disorders, including IBD; BACH2 affects intestinal tight junction formation ^52–57^. Since the parent with UC and Crohn’s disease does not carry the *BACH2* variant, an oligogenic bilineal inheritance pattern is likely.

### STRING

To explore functional relationships among the gut genes described in this paper, we performed a protein–protein interaction (PPI) analysis using the STRING database (version 12.0; https://string-db.org/) (**Figure**). The candidate genes were separated into three different functional categories related to gut physiology and gut immune function: SLC10A2, LGALS4, and MEP1B overlapped in their antigen processing function and luminal interface localization; DUOX2, LGALS4, and TTC7 affect oxidative stress and antimicrobial defense, and BACH2 and CCR9 are involved in intestinal immune function.

**Figure.**
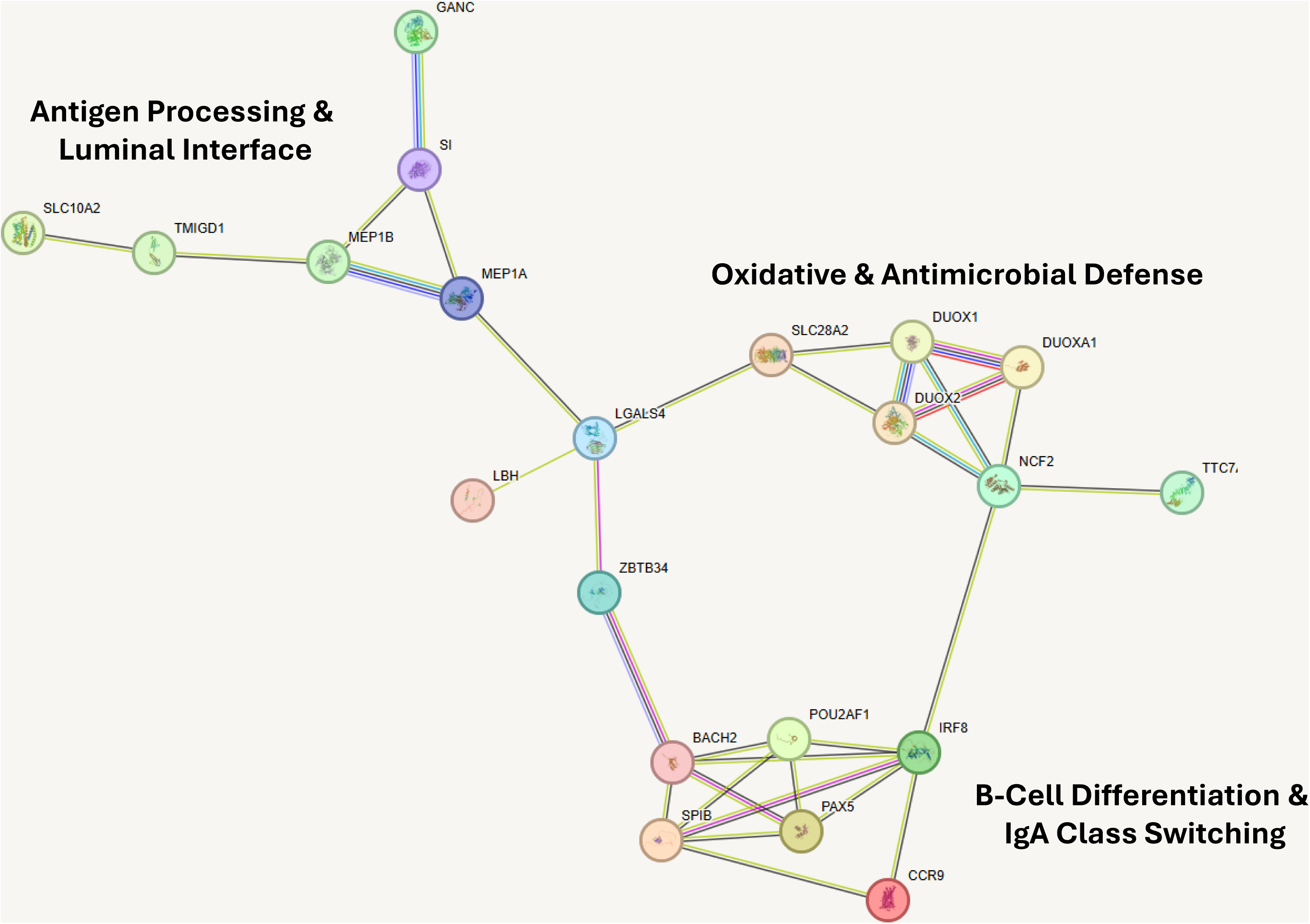
STRING: Protein-Protein Interaction Network showing separation of gut candidate genes into distinct functional categories

## Discussion

We have identified nine genes with pathogenic or likely pathogenic mutations in patients with PANS and/or regression in ASD that are expressed primarily in the gut and are strongly associated with Crohn’s disease, intestinal permeability, and/or gut dysbiosis. Gut dysbiosis is a disturbance in the balance of the microbial ecosystem that may contribute to neuropsychiatric and neurodegenerative disorders through effects on the gut-immune-brain axis ^58–62^. This is a bidirectional communication pathway that connects the brain, intestines, and intestinal microbiota via the autonomic nervous system through the vagus nerve, and neuroendocrine, neurotransmitter, and neuroimmune pathways ^63–65^. Gut dysbiosis can disrupt the gut barrier (intestinal permeability), thereby promoting inflammation and innate immune activation via toll-like receptor signaling by pathogenic microbes or their metabolites, such as lipopolysaccharide, peptidoglycans, branched-chain amino acids, flagellin, indole derivatives, and secondary bile acids ^62, 66–68^.

The gut is also an organ of the immune system containing approximately three-quarters of the body’s immune cells and plays a role in protecting against infectious microbes and the development of autoimmune disorders ^69–71^. Commensal gut microbes influence mucosal immunity and shape the development and regulation of systemic immune responses ^72^. However, the underlying molecular and microbiological pathogenesis of neuropsychiatric symptoms related to the gut are poorly understood. Based on the findings in this paper, genetic factors are playing a role.

The most striking candidate gene we identified is *DUOX2*, with ultrarare pathogenic or likely pathogenic variants (MAF 0.00008 to 0.0002) occurring in four cases. The fact that two cases were identified in each of the cohorts adds strength to the finding. While our cohorts are too small for formal statistical analysis, the presence of these variants in over 3% of the included patients warrants further investigation. However, it must be pointed out that there is some ascertainment bias regarding the *DUOX2* gene since the parents of one case introduced the corresponding author to another case.

*DUOX2* codes for an enzyme that has a crucial role in producing hydrogen peroxide and thyroid hormones. As noted above, it is expressed at the apical membrane of gastrointestinal epithelia and contributes to IBD by disrupting microbiota-immune homeostasis ^18, 19^. Single-cell RNA sequencing in Crohn’s disease showed that *DUOX2* is highly expressed in distinct epithelial cell types in both the terminal ileum and ascending colon ^73^, and was found to be a top differentially expressed gene compared to healthy controls ^74^.

Finally, exome sequencing in two large cohorts identified a functional nonsynonymous *DUOX2* variant in Crohn’s disease (P303R) ^75^. *DUOX2* has also been implicated in ulcerative colitis; it is one of six hub genes that were found to be differentially expressed in a mouse model ^76^. *DUOX2* expression is also a biomarker associated with ulcerative colitis and rheumatoid arthritis in RNA sequencing studies ^77, 78^. These findings show a strong association between DUOX2 and IBD. In the context of PANS, it is particularly interesting that *DUOX2* expression interacts with both the proinflammatory cytokine IL-17 and gut barrier immunity ^21, 22^. IL-17 has been implicated in psoriasis, which is a frequent comorbidity in PANS families ^79^. Also, IL-17 has been shown to be involved in the pathogenesis of post-streptococcal PANS (PANDAS: Pediatric Autoimmune Neuropsychiatric Disorders Associated with Streptococcal Infections) in a mouse model ^80^.

In addition to *DUOX2*, we found several other candidate genes that are associated with gut dysbiosis, IBD, and/or intestinal permeability. *SLC10A2,* for example, codes for the apical sodium-dependent bile acid transporter (ASBT) that is expressed almost exclusively in the small intestine and plays a key role in the reabsorption of bile acids ^81^. Bile acids are steroids formed at the interface between host metabolism and intestinal microbiota, with primary bile acids generated in the liver and secondary bile acids produced by microbial enzymes ^82^. Bile acids modulate intestinal permeability where disrupted metabolism due to dysbiosis can impair gut barrier function and exacerbate inflammation in humans and animal models ^83, 84^.

Bile acids also modulate immune responses by binding to bile acid receptors, thereby modulating immune responses and inflammatory reactions, including the balance between Th17 and Treg cells, blood-brain barrier integrity, and the interaction of peripheral immune cells with the brain ^85, 86^. The effect on Th17 cells or IL-17 may be a common mechanism linking *SLC10A2* to *DUOX2* and several other gut candidate genes in PANS; *BACH2, LGAL4*, and *CCR9* are other genes in this study that have connections to IL-17 and Th17 function affecting autoimmunity and intestinal inflammation ^53, 87, 88^. In addition, hydrogen peroxide, which is produced by the action of DUOX2, significantly affects bile acids and their functions ^89^.

Case 5, the sibling of Case 5, and Case 6 all have PANS and carry a pathogenic *SLC10A2* variant.

Case 6 also developed severe spondylarthritis in grade school and the Case 5 sibling has been diagnosed with PFAPA. We have previously identified a PANS case who was also diagnosed with PFAPA, as well as a sibling pair with Familial Mediterranean Fever (FMF) ^6^. Physicians caring for FMF and PFAPA children should be aware of these interactions and consider the possibility that neuropsychiatric issues could have an immune/inflammatory-based etiology requiring unique diagnostic and treatment strategies.

*LGALS4* codes for galectin-4, a member of the galectin family of beta-galactoside-binding proteins implicated in modulating cell-cell and cell-matrix interactions ^90^. This gene is expressed almost exclusively in epithelial cells in the transverse colon and terminal ileum. LGALS4 and other galectins are involved in the development of IBD by regulating T-cell function in the gut and affecting host-gut microbe interactions ^40, 41^. Expression of common mucosal-associated galectins, including galectin-4, is dysregulated in inflamed tissues of IBD patients compared to controls. LGALS4 binds to intestinal epithelial cells and promotes their restitution, playing a significant role in intestinal wound-healing that occurs in IBD ^91^.

SI (sucrose-isomaltase) deficiency is a cause of diarrhea-predominant irritable bowel syndrome; a condition associated with dysbiosis ^42, 43^. SI is a marker for intestinal epithelial cell differentiation that is decreased in inflamed regions in a mouse IBD model ^92^.

Case 11 has a de novo frameshift mutation in *CCR9*, which codes for the CCL25 chemokine receptor. CCL25 is a chemokine that affects gut homeostasis and permeability ^44–46^. Its expression increases in colitis and correlates with inflammatory activity ^93^. CCL25 binds to CCR9 on immune cells, guiding them to the gut lining. This is a critical part of the immune response that maintains gut health, but it becomes problematic in inflammatory conditions. CCR9/CCL25 signaling is implicated in autoimmune and autoinflammatory disorders beyond IBD, including autoimmune pancreatitis, SLE, and Sjögren’s Syndrome ^94, 95^. The effect of a signaling pathway expressed primarily in the intestines on autoimmune and autoinflammatory disorders points to the interaction between the gut and systemic immune systems ^96, 97^. Finally, in a randomized, double-blind, placebo-controlled trial of CCR9-targeted leukapheresis, a therapeutic effect on ulcerative colitis was observed^98^. Like CCR9, CCL25 is almost exclusively expressed in the intestines ^99^. CCR9 may also impact Th17 cell numbers and function in IBD and other inflammatory disorders ^100^.

*MEP1B* codes for a metalloprotease that plays a role in the hydrolysis of proteins that have been implicated in cancer, intestinal inflammation, and ECM remodeling ^50^. It is almost exclusively expressed in the small intestine and duodenum. MEP1B and other meprins have been implicated in IBD ^101, 102^. MEP1B also plays a role in regulating intestinal permeability by promoting mucus detachment, which helps prevent bacterial overgrowth ^50^. Gene expression analysis in Crohn’s disease vs controls showed that *MEP1B* was among the top downregulated genes ^103^. On the other hand, *MEP1B* was among the top upregulated genes in ulcerative colitis ^104^.

Case 7 has a pathogenic mutation in *TTC7A*, which codes for tetratricopeptide repeat domain 7A. Homozygosity or compound heterozygosity for loss-of-function mutations causes a profound disruption of the epithelial barrier along the entire gastrointestinal tract causing Multiple Intestinal Atresias (MIA) combined with Combined Immunodeficiency (CID: MIA-CID), and tricho-hepato-enteric syndrome (SD-THE) IBD ^37–39^. The effect of heterozygosity for a pathogenic variant is unclear. However, a sibling pair with early-onset IBD was found with a nonsynonymous mutation in *TTC7A* in one study. suggesting that heterozygosity can have clinical consequences, although a hidden effect of the other allele through mutations in non-coding DNA cannot be ruled out ^105^. Case 7, his sibling and the sibling of case 1 also harbor a common *NOD2* variant that has been associated with common variable immune deficiency (CVID), Blau syndrome, and Yao syndrome ^30–34^. *NOD2* common variants have been found in Crohn’s disease in many case-control association studies ^106–108^. Case 7 is interesting because his sibling has celiac disease and has the same *TTC7A* and *NOD2* mutations. Although she does not meet clinical criteria for PANS or regression, she does have dysautonomia and hypermobility, which are commonly found in PANS ^109^

Case 12 and his siblings, along with their parent who has Crohn’s disease, have a pathogenic (predicted by AM) mutation in *UNC45A,* which codes for a protein that regulates intestinal epithelial barrier integrity and repair. The finding of another *UNC45A* variant in case 4 that was scored as pathogenic by AM supports a role for this gene in PANS and regression.

*BACH2* is a ubiquitously expressed basic leucine zipper transcription factor that plays a crucial role in regulating Treg and B cell formation ^52–54^. Case 13, who has a de novo *BACH2* mutation, has low IgG/IgM levels and markedly reduced B cells, which is consistent with a diagnosis of BACH2-related immunodeficiency and autoimmunity (BRIDA) ^110, 111^. *BACH2* has been linked to IBD at both the molecular and genetic levels ^55–57^. Heterozygous mutations in the *BACH2* gene increase susceptibility to other autoimmune and autoinflammatory disorders ^112^. Interestingly, a GWAS study evaluating the presence of thyroid peroxidase antibodies, a risk factor for Hashimoto’s thyroiditis and Graves’ disease, showed an association with a *BACH2* single nucleotide polymorphism ^113^. While the *BACH2* variant in case 13 is reported as *de novo*, this requires Sanger validation. The paternal history of ulcerative colitis raises questions regarding multifactorial inheritance or shared environmental triggers. Given the known link between ulcerative colitis and dysbiosis, it remains to be determined whether the transmission of specific microbial species between family members might act as a secondary ’hit’ in the context of a *BACH2* mutation ^114, 115^.

Eight out of 13 cases had secondary mutations of interest in the context of innate immunity, DNA repair, or IBD, as noted for NOD2 above. Others include *STK19* and *UVSSA* in case 1; CFI in case 3; *USP45* and *PARP14* in case 10; and *NLRC4* (case 2)*, UNC45A* (case 4)*, LRRC8A* (case 5), and *PRKN* (case 8) (**Supplementary Table S1**). The roles of *STK19/UVSSA, USP45/PARP14,* and *PRKN* in the neuropsychiatric manifestations of PANS and regression, in the context of DNA repair and mitochondrial function, have been described in another paper ^6^. Among these secondary genes, *NOD2, CFI, PARP14*, and *NLRC4* have been linked to Crohn’s disease at the molecular or genetic level, or in animal models ^49, 116–120^. *NLRC4* codes for a component of the inflammasome, a cytosolic multiprotein complex that assembles in response to exogenous or endogenous stressors that play a significant role in autoinflammatory diseases, macrophage activation syndrome, and panoptosis ^79, 121^. *LRRC8A* regulates NLRP3 inflammasomes along with other genes coding for ion channels and transporters involved in immune cell development and function ^122, 123^. CFI, also known as C3b/C4b inactivator, could result in activation of the complement cascade if the mutation is a loss-of-function variant. CFI deficiency is a rare autosomal recessive disorder that impairs the regulation of the complement system. This leads to increased susceptibility to infections, particularly those caused by encapsulated bacteria, and is also associated with autoimmune diseases, including Acute Disseminated Encephalomyelitis and SLE ^124, 125^.

All patients had severe gastrointestinal symptoms, as reported by their parents, with the possible exception of case 4 who instead repeatedly experienced nausea and vomiting following Streptococcus pharyngitis (**Table 2**). Symptoms are heterogeneous, including chronic abdominal pain, diarrhea, constipation, gastroesophageal reflux, and reported food allergies to gluten and dairy products. In case 13, severe gut symptoms preceded catatonia relapses. A minority of cases had an extensive GI evaluation (cases 1 and 7). However, because many children with PANS and ASD/NDD complain of intestinal problems, it is not clear if the GI symptoms found in our small cohort are related to the genetic variants we found or are an epiphenomenon.

Co-existing diagnoses of PANS and IBD have previously been described by Tang et al. (Crohn’s n=5, ulcerative colitis n=3) ^79^. In seven of the eight cases, PANS predated the diagnosis of IBD by 1-15 years. In one case, however, IBD occurred 8 years before the first PANS flare. Interestingly, there was a tendency for PANS flares to improve with treatment of IBD. In our cohort, patients had GI symptoms, but IBD has not been diagnosed in any case yet. However, there is a positive family history of IBD in cases 4, 12, 13. In case 1, GI symptoms, as well as other clinical features (e.g., enthesitis and arthritis), improved following treatment with Humira. In case 13, severe gut symptoms preceded catatonia relapses. Future research is required to confirm if the observed correlation between GI symptom resolution and psychiatric recovery indicates a causal gut-brain link or if these findings are simply anecdotal.

A large body of research suggests involvement of dysbiosis and intestinal permeability dysfunction, and neuropsychiatric and neurodegenerative disorders ^126^. However, a comprehensive review by Mitchell et al. (2026) disputes this conclusion ^127^. Mitchel et al. contend that small sample sizes, potential type I errors, and various methodological limitations in both murine and human research preclude a definitive link between the gut microbiome and the pathogenesis of ASD. Despite the difficulty in drawing universal conclusions, the findings reported here suggest that gut microbial factors may play a role in a proportion of patients with specific genetic predispositions.

In addition to the gut genes described in this paper, several cases also harbored ultrarare mutations in DNA repair and immunological pathways, possibly amplifying immunogenetic vulnerability. However, it should be noted that the small sample size and ascertainment bias are weaknesses of our study. The lack of whole genome or whole exome sequencing in a majority of cases is also a weakness. Other problems include reliance on non-structured family histories, retrospectively reported gastrointestinal symptoms in some cases, and incomplete gastrointestinal evaluations. Nevertheless, the findings are of potential interest as many of the candidate genes are also identified as candidate risk genes for autoinflammatory or autoimmune diseases providing clues toward mechanisms and individualized treatment strategies. The connection between genetic variants that affect gut homeostasis and acute neuropsychiatric decompensation in this small study sample will require pathophysiological studies using human gut organoids and murine models. Advancing this line of inquiry may facilitate more accurate patient stratification and the development of targeted therapies, such as gut permeability repair or fecal microbiota transplantation, for specific subgroups of patients with PANS and regressive symptoms.

## Methods

### Ethics Disclosure

This study was conducted in accordance with the Declaration of Helsinki. Informed consent was obtained from all subjects and/or their legal guardian(s). The study was approved by the Albert Einstein College of Medicine and Stanford University Institutional Review Boards (IRB). All experiments were performed in accordance with IRB guidelines and regulations.

### Subjects

For the Einstein cohort,105 patients were identified through connections between the senior investigators and a PANS group called EXPAND (https://expand.care/), the Neuroimmune Foundation (https://neuroimmune.org/about/), the Alex Manfull Fund (https://thealexmanfullfund.org/), and the Stanford Immune Behavorial Health clinic (who request consultation from H.M.L on select cases which are included in his broader studies). Histories were obtained by contributing clinicians and confirmed and collated by one of the senior investigators (H.M.L.). The cases met criteria for PANS or had other regressions/suspected neuroimmune deteriorations with or without comorbid ASD or NDD. In cases with acute or subacute onset of behavioral regression, caregivers and clinicians observed that the loss of previously established skills (e.g., speech, math/reading skills, handwriting/drawing, executive function, urinary continence) and daily functioning. Each patient was assigned to a study ID known only to the research group, thereby maintaining patient confidentiality. In addition to the Einstein cohort, 23 cases who had been analyzed using Invitae panels were obtained from the Stanford Immune Behavioral Health Clinic. Subject selection from this cohort is described in **Supplementary Figure S2**. WES is currently being carried out in the 301 PANS cases from which the 23 ascertained in this study that were derived.

### Genetic analysis: Variant calling and annotation

The senior author has assembled a dataset utilizing WES (n=36) and Invitae panels (n=69). Data from WES analyses were processed in the Computational Genomics Core at the Albert Einstein College of Medicine after importing sequencing files from GeneDx, Nebula, and CeGaT. Commercially generated VCF files that had been mapped to the human genome (hg38) were annotated with Annovar (version: Date: 2020-06-08) ^128^; commercially generated VCF files that had been mapped to hg19 were re-mapped to hg38 using the Picard Tools module LiftoverVcf (version: 2.26.10) before annotation. In cases where LiftoverVcf files were not available, FastQ files were aligned to hg38 using the MEM algorithm of BWA (version 0.7.17-r1188; https://bio-bwa.sourceforge.net), and bam files were sorted by coordinates, duplicates marked, and base quality scores were recalibrated (Picard Tools modules SortSam and MarkDuplicates; Genome Analysis Toolkit (GATK) v4.4.0.0 modules BaseRecalibrator and ApplyBQSR). Variant calling was performed using Haplotypecaller and raw variants were filtered using modules CNNScoreVariants (1D model, pre-trained architecture) and FilterVariantTranches (GATK, v4.4.0.0); filtered VCF files were annotated by Annovar. In all cases, only entries that passed all filters (PASS in the VCF file FILTER column) were annotated. QC was performed using FastQC, FastQ-screen, and CollectHsMetrics.

For this current study, genes of interest included ultrarare pathogenic variants that were both expressed in the gut and associated with dysbiosis, altered intestinal permeability, and/or inflammatory bowel disease and were identified in the above larger data sets: WES (n=6) and Invitae (n=7). All ultrarare variants (minor allele frequency (MAF) <0.001) in exons and intron/exon junctions were assessed. MAFs were obtained from the GnomAD database on the UCSC Genome Browser (https://genome.ucsc.edu/). We focused on splice variants, frameshift and non-frameshift mutations, stop-gain and stop-loss variants, and indels. In addition, all ultrarare nonsynonymous variants in ASD and NDD-related genes identified in the SFARI database and in Invitae NDD panels (Immunodeficiency and Autoinflammatory, Autoimmunity Syndromes, DNA Damage Repair, and Nuclear Mitochondrial Disorders) predicted initially to be pathogenic by SIFT and MutationTaster were also evaluated (see the next section).

### Evaluation of nonsynonymous variants

The predicted pathogenicity of each variant, based on ACMG criteria (pathogenic, likely pathogenic, uncertain, possibly benign, likely benign), was obtained from the Franklin database and, in some cases, from ClinVar and VarSome; a few exceptions will be described on a case-by-case basis.

https://franklin.genoox.com/

https://www.ncbi.nlm.nih.gov/clinvar/

https://help.genoox.com/en/articles/6240723-prediction-tools-pp3-bp4

https://varsome.com/

We also used CADD scores in assessing single nucleotide variants. CADD provides a Phred-like score that is a measure of deleteriousness ^129^. A CADD score represents a ranking, not a prediction, and no threshold is defined. Higher scores are more likely to be deleterious. A score of 20 is commonly used as a cutoff, with scores above 20 indicating that a substitution is among the 1.0% most deleterious possible substitutions in the human genome. CADD scores were obtained using a tool developed by the University of Washington (https://cadd.gs.washington.edu).

Finally, nonsynonymous mutations were also evaluated using AlphaMissense (AM), an adaptation of AlphaFold that evaluates amino acid changes to predict the pathogenicity of nonsynonymous variants. (Cheng et al., 2023). It combines the structural context of the amino acid substitution and evolutionary conservation to provide a pathogenicity score. AM predicts the pathogenicity of all possible single amino acid substitutions across nearly all protein-coding genes, enabling classification of 89% of nonsynonymous variants. The variants are scored as pathogenic, ambiguous, or benign based on the score, with ambiguous falling just outside the pathogenic range. Finally, SpliceAI was used to evaluate splice site variants. A score of 0.5 (recommended) was used as a cutoff (https://spliceailookup.broadinstitute.org/).

Several candidate mutations were validated by Sanger sequencing (**Supplementary Text S3**).

### Protein–Protein Interaction Network Analysis

The online available STRING (Search Tool for the Retrieval of Interacting Genes/Proteins is a biological database and web resource of known and predicted protein–protein interactions.

This resource was used in combination with the Human Gene Mutation Database (HGMD®) which represents a repository that contains known (published) gene lesions responsible for human inherited diseases. The HGMD repository describes functionally proven inherited mutations useful for medical research, genetic diagnosis, and next-generation sequencing studies providing candidate variants linked to gut biology diseases and behavioral related phenotypes. The interaction network describes the biological relevance of the identified patients in relation to the underlying molecular pathways that are affected in these patients.

### Expression pattern

The expression pattern of each candidate gene was assessed on the UCSC Genome Browser (https://genome.ucsc.edu) and the Human Protein Atlas (https://www.proteinatlas.org/).

## Data Availability

All data produced in the present work are contained in the manuscript

## Acknowledgments

The authors want to thank the participating families.

## Author contributions

H.M.L. designed the study, co-wrote the manuscript, processed WES, interviewed families, and designed and conceived the study; J.F. co-wrote the manuscript and contributed cases; R.A.B. processed WES, J.L.C. co-wrote the manuscript; E.P. processed DNA, validated variants, edited manuscript; E.S and N.R. contributed cases; A.S. validated variants; D.V. validated variants; P.J.S. carried out STRING analysis; C.N. and N.S. contributed cases with J.F, and N.S. edited the manuscript. All authors reviewed the manuscript.

## Data availability

Not applicable.

## Supplementary Information

**Supplementary Table S1:** Secondary genes of interest. Abbreviations as in Table legend

**Supplementary Figure S2.** Ascertainment flow chart for the Stanford cohort

**Supplementary Text S3**. Method for validating selected candidate genes

