## Supplemental Table 1 for "Ultrarare Variants in Genes Involved in Intestinal Microbiota and Permeability Homeostasis in Youth with Developmental and Neuropsychiatric Deteriorations"

| Case# | Study ID | Diagnosis |
|---|---|---|
| 1 | DUZG | PANS |
| 2 | DUMO | PANS |
| 3 | DUWJ | PANS |
| 4 | DUHU | PANS |
| 5 | OC_PANS | PANS/catatonia |
| 6 | 77-0403 | PANS |
| 7 | EZ_PANS | PANS |
| 8 | KDWP | ASD/PANS |
| 9 | EH_PANS | PANS; chronic bulimia and depression |
| 10 | MKJ_PANS | PANS |
| 11 | DAJ_PANS | ASD/verbal apraxia/PANS |
| 12 | CAS_PANS | PANS |
| 13 | CO_AE | ASD, catatonia, AE |

| Secondary gene | Variant |
|---|---|
| STK19, UVSSA | splice donor, c.815+1G>A/splice acceptor, c.551-1G>T |
| NLRC4 | NLRC4 (c.1276C>T p.Leu426Phe) |
| CFI | c.848A>G, p.Asp283Gly |
| UNC45A | c.2003C>A, p.Ser668Tyr |
| LRRC8A | c325G>T, p.Val109LIeu |
| NONE | |
| NOD2 | c.2104C>T, Arg702Trp |
| PRKN | exon 2/3 del |
| NONE | |
| USP45/PARP14 | stopgain, A1462T; p.K488*/splice donor exon2:c.321+1G>A |
| NONE | |
| NONE | |
| NONE | |

| rs # or chr. pos. | MAF |
|---|---|
| rs752938149/rs886579502 | 0.0001/0.000014 |
| chr2:32250588 | Novel |
| rs756201106 | 0.000015 |
| rs144806267 | 0.0006 |
| rs370697938 | 0.00006 |
| rs2066844 | 0.02 |
| Chr6:162643775-162940281 | unknown |
| rs189281869/rs201755391 | 0.0001/0.0008 |

| CADD/AM | SpliceAI score |
|---|---|
| 33/33 | 0.54/0.95 |
| Unknown/ambiguous | N/A |
| 24.3/pathogenic | NA |
| 28.3/pathogenic | NA |
| <5/psthogenic | NA |
| 23.1/NA | NA |
| N/A | NA |
| 39/NA | NA |
| 34/NA | N/A/0.71 |
| 36/NA | NA |
| NA | NA |

| Franklin | transmision |
| --- | --- |
| VUS/likely pathogenic | both paternal |
| VUS | unknown |
| likely pathogenic | unknown |
| VUS | maternal |
| VUS | unknown |
| benign | unknown |
| N/A | de novo |
| pathogenic | maternal |
| likely pathogenic/ VUS | maternal/paternal |
| VUS | unknown |
| VUS | de novo |
