## Supplemental Figure for "Ultrarare Variants in Genes Involved in Intestinal Microbiota and Permeability Homeostasis in Youth with Developmental and Neuropsychiatric Deteriorations"

### Supplementary Figure S2: Subject selection (Stanford Immune Behavioral Health Clinic)

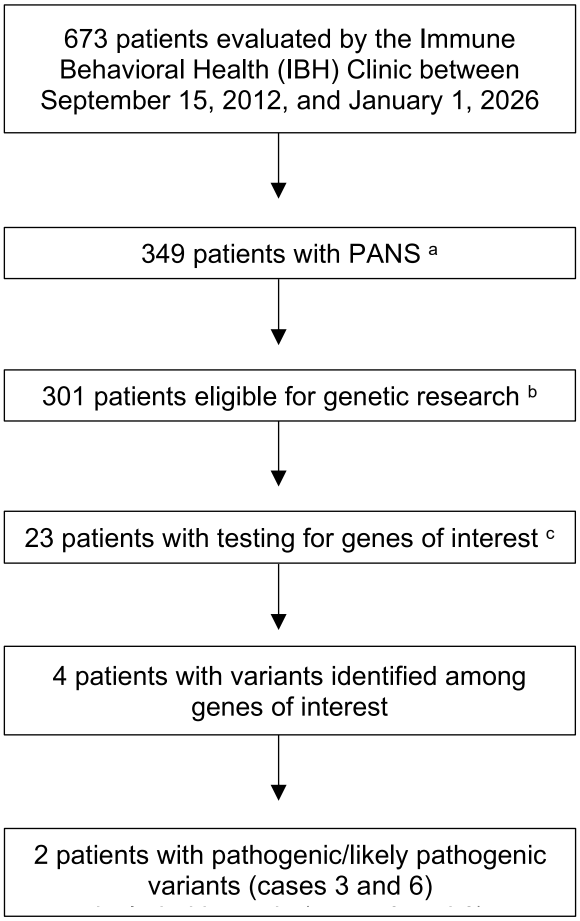


[a] Patients were diagnosed with PANS according to diagnostic criteria (Chang et al. 2015).
[b] Patients were considered eligible if they or a parent (in the case of a minor) gave informed written consent for genetic research. Competent patients aged 7-17 must also provide assent.
[c] Genes of interest are *DUOX2*, *SLC10A2*, *BACH2*, *SI*, and *TTC7A*. 21 of 23 subjects were tested for all genes of interest. 2 subjects were tested for all genes except *SLC10A2*.
