## Supplemental Methods for "Ultrarare Variants in Genes Involved in Intestinal Microbiota and Permeability Homeostasis in Youth with Developmental and Neuropsychiatric Deteriorations"

### Supplementary Methods

#### Validation of selected variants

Several variants were validated by Sanger sequencing using saliva DNA, which was obtained using a DNA Genotek kit (cat# OGR-500). DNA was extracted using prepIT purifier (DNAgenotek cat# PT-L2P-5) according to the manufacturer's instructions, except for the inclusion of a phenol/chloroform purification step (Phenol:Chloroform:Isoamyl Alcohol 25:24:1 saturated with 10mMTris pH8.0, 1mM EDTA; followed by the addition of 1ul glycogen, sodium acetate, pH 5.2 to a final concentration of 300 mM, and an equal volume of Isoamyl Alcohol to precipitate DNA). Genomic DNA was amplified using HotStarTaq DNA Polymerase (Qiagen cat# 203203) according to the manufacturer's instructions using primers that flank the variants of interest (see list below). PCR products were purified using a QIAquick PCR Purification Kit (Qiagen, catalog# 28104) according to the manufacturer's instructions. DNA was sequenced by the standard Sanger dideoxy chain termination method using one of the two PCR primers. The primers used are shown below.

DUOX2-F: CGTCACCTGGTTGGCCTG
DUOX2-R: TTCCTCAACATCCGCATCCC

SI-F: GTAACATGCCTTTTGATCATTTTACT
SI-R: ATAAGAGAAGCTTCATTCTTACCCG

CCR9-F: CCCTTGCAGAGCCCTATTCC
CCR9-R: AGAGGAGGTCAGCAATTGCC

STK19-F: TGCGCTCAGATCAAGAATCCA
STK19-R: GCGGATTGGCTCCCTCCA

USP45-F: TAAGACCTCAAATGCTTCCC
USP45-R: GGTGATAAGGAAATGGCAGA

MEP1B-F: AGGTGATCCACATCTTTAACTGTGA
MEP1B-R: TGCGGAGTCAAATTTGGTCG
